# VARIANCE: Variability of Assessed BP Readings In Acute Neuro-intervention Cases and its Effect on outcomes

**DOI:** 10.1101/2023.12.05.23299576

**Authors:** Kimberlee Van Orden, Thomas Staniszewski, Maryo Jajo, Dolores Torres, Briana Poynor, Benjamin Alwood, Kunal Agrawal, Brett Meyer, Dawn Meyer

## Abstract

**Background:** Blood pressure variability (BPV) following endovascular thrombectomy (EVT) in AIS has been associated with poor outcome. Variables that predict high BPV must be examined to improve outcome. The purpose of this study is to analyze predictors and effect of BPV on outcome in patients with good recanalization after EVT.

**Methods:** We conducted a retrospective analysis of prospectively collected data from an IRB approved registry of two academic Comprehensive Stroke Centers between 2017-2022. Patients were included if they had 1) anterior circulation large vessel occlusions (LVO) and 2) EVT with TICI 2b-3. All BPs were recorded as immediate pre-op SBP/DBP (one minute prior to procedure), immediate post-op SBP/DBP (at time of recanalization), and 24 hours post revascularization. Demographic variables, stroke time metrics, and symptomatic ICH (sICH) were assessed. Outcome included discharge disposition and 90 day modified Rankin Scale. Good short-term outcome was disposition to home/acute rehabilitation and good long-term outcome was mRS 0-2. BPV was the average of the differences between measurements divided by the number of measurements. R

**Results:** We identified 253 patients (mean age 70±14, 51.4% female, 49% white, 30.4% Hispanic). Median NIHSS was 17±8 and mean onset to groin puncture was 478±326 mins. Mean door to groin puncture was 86±110 mins. Good discharge occurred in 58.9% of patients. Mean BPV was 30.6±25.6mmHg and was significantly correlated with female sex, home antihypertensive use, and immediate pre and post-op SBP. BPV was not associated with age, initial NIHSS, Hispanic ethnicity, HTN, sICH, discharge disposition or 90 day mRS.

**Conclusion:** There was a significant correlation of high BPV with female sex and home antihypertensives use. BPV was not correlated with short or long term outcome in this population, however, BPV was lower than prior populations. Assessment of BPV in various stroke centers and populations is necessary to understand the effect of BPV on stroke outcome.

## Introduction

Blood pressure variability (BPV) is defined as the continuous and dynamic fluctuations that occur in blood pressure levels throughout a specific time period^1^. BPV is practically examined as the ranges from high to low systolic and diastolic measurements. High BPV occurs when there are wide ranges from highest and lowest blood pressure values. Low BPV occurs when there is a narrow range^2^.

Studies support that increased BPV in the hyperacute period of acute ischemic stroke (AIS) may have more impact on overall hemorrhage risk and outcome as opposed to the mean blood pressure levels^3^. Clinical studies have identified a significant relationship between BPV and outcome in both the short term up to hospital discharge and long term at 90 days^4,5^. A systematic review and meta-analysis found that reducing BPV in AIS patients was associated with improved functional outcomes and reduced mortality^6^. The association has been shown to be more robust for systolic BPV compared to diastolic BPV in the stroke population^2^.

Evidence based recommendations have been well-established for blood pressure (BP) targets in AIS patients receiving acute thrombolytics^7^. While guidelines do exist for BP goals in AIS patients with large vessel occlusive (LVO), the data is much less robust and large, well designed and executed randomized trials are lacking^8^. Stroke patients with LVO are more likely to have a large penumbra and this penumbra can be more susceptible to the negative effects from fluctuating cerebral perfusion with high BPV^9^. High BPV has also been shown to increase the rate of symptomatic intracranial hemorrhage (sICH)^10^. It is critical to establish the relationship of BPV in LVO in order to optimize outcomes and reduce the risk of sICH. High BPV following endovascular thrombectomy (EVT) in AIS patients has been associated with poor functional outcomes both in the short and long term^8^. Short term functional outcome at hospital discharge disposition has been shown to be worse in LVO patients treated with EVT when controlling for mediating factors^11^. A post hoc analysis of BEST (Blood Pressure after Endovascular Therapy for Ischemic Stroke) demonstrated that higher BPV in the first 24 hours is associated with worse 90-day outcome^8^. Establishing targets for both SBP and DBP goals in addition to BPV targets may provide the best opportunity for optimal functional outcome with reduced risk in the post-EVT population.

Identifying mediating factors and outcomes in BPV are critical to develop interventions and improve stroke outcome. The role of sex differences and degree of recanalization in acute stroke therapies must be examined. The purpose of this study was to assess BPV and its impact on short and long term outcome in AIS patients who underwent EVT with successful recanalization. Sex differences in BPV were also examined.

## Methods

### Study population

Data was retrospectively examined from a prospectively collected, IRB approved Stroke Registry at two academic Joint Commission (TJC) accredited Comprehensive Stroke Centers in Southern California between 2017-2022. The registry includes all patients for whom a stroke code is activated at the CSC. Subjects were included if they had: 1) focal neurologic deficit; 2) anterior circulation large vessel occlusion (LVO) including MCA, ACA, ICA; 3) and had endovascular treatment (EVT) with TICI 2b-3 recanalization.

### Blood Pressure Measurement and Blood Pressure Variability

All BPs were recorded as immediate pre-op SBP and DBP (one minute prior to procedure start time), immediate post-op SBP and DBP (at time of recanalization), and 24 hours post revascularization. All measurements were done with non-invasive monitoring.

BPV was defined by the average real variability (ARV) calculated as the absolute value of the average of the differences between consecutive BP measurements divided by the number of measurements. BPV was calculated by the average real variability (ARV) calculated as the absolute value of the average of the differences between consecutive BP measurements divided by the number of measurements.

### Study Outcomes

The primary outcome of the study was short term outcome as defined by hospital discharge disposition. Good disposition was defined as hospital discharge to either home or an acute rehabilitation facility (ARU). Poor disposition was defined as discharge to skilled nursing facility (SNF), death, Hospice, or other. Secondary outcome was defined as modified Rankin Scale (mRS) at 90 days. Good long-term outcome was mRS of 0-2 and poor as mRS 3-6.

### Statistical Analysis

Variables examined include age, sex, Hispanic ethnicity, initial NIHSS, history of HTN, current antihypertensive use, symptom onset to groin puncture time, door to groin puncture time, and symptomatic ICH (sICH). Male and female subjects were compared for age, sex,Hispanic ethnicity, initial NIHSS, history of HTN, current antihypertensive use, symptom onset to groin puncture time,door to groin puncture time, and symptomatic ICH (sICH), BPV, onset to groin, and door to groin.

A correlation matrix was used to identify both continuous and categorical variables with a significant correlation with ARV (p≤0.10). A stepwise logistic regression model for good versus poor outcome was created by including all significant variable from the correlation matrix. A significance level of p≤0.05 was included for all non-correlational analysis.

## Results

We identified 253 patients (mean age 70±14 years, 51.4% female). 49% were white, 6.3% Asian, 5.5% black, 1.6% native Hawaiian pacific islander, and 37.5% are unspecified. 30.4% were Hispanic. Median NIHSS was 17±8. Mean BMI was 27kg±8kg. Mean symptom onset to groin puncture time was 478±326 mins. Mean door to groin puncture time was 86±110 mins. 34.8% of patients received tPA (100% of those eligible as our population included patients in the extended window who qualified per DAWN/DEFUSE-3). 57.2% had cardioembolic etiology. 4.7% had sICH. 58.9% of patients had a good discharge disposition.

Overall BPV, defined as the average SBP variability as an absolute value, was 30.6±25.6. BPV was associated with sex, home antihypertensive use, and immediate pre-op and post-op SBPs. BPV was not associated with age, initial NIHSS, Hispanic ethnicity, HTN, rate of sICH, or short or long term outcome.

For sex differences assessment, we included 131 females and 124 males of the overall sample. There was a significant difference between males and females in: 1) history of a fib (76% v 64%, p=0.42); 2) atrial fibrillation during admission (36% v 50%, p=0.01); 3) age (male 67 v female 73 years, p<0.001); 4) post-EVT SBP (137mmHg v 144mmHg, p-0.02). There was a significant difference in onset to groin (p=0.41), but no significant difference in door to groin (p=0.59). When adjusting for time of onset, there was no significant difference in thrombolysis prior to EVT between groups. There was no significant difference in sICH (6 male v 4% female, p=0.50), 90 day mRS (p=0.81); or discharge disposition (p=0.56). BPV was significantly correlated with sex but not significant for any interaction effect with the variables assessed.

## Discussion

The mechanisms underlying the detrimental effects of BPV on acute stroke outcomes are multifactorial. Fluctuations in BP can lead to impaired cerebral autoregulation, resulting in inadequate perfusion and ischemic injury to the brain (ref). It is possible that repeated episodes of hypoperfusion and reperfusion due to BPV may exacerbate tissue damage and hinder the recovery process.

The purpose of this study was to determine the impact of BPV on short and long term outcome in stroke patients undergoing successful EVT. There was a significant correlation with female sex, age, and home use of antihypertensive medications and higher BPV. These variables can potentially be used as predictors of higher BPV and allow for identification of subgroups at higher risk of high BPV.

BPV was not correlated with discharge disposition or 90 day mRS in our population. We hypothesize that this was due to the low variability in this study compared to previous studies. Prior research has shown that BPV is independently associated with the development of neurologic deterioration in the acute stage of ischemic stroke (ref) and that early neurologic deterioration is correlated with poor outcomes (ref). However, these studies did not specifically examine acute stroke patients post EVT with successful reperfusion. Post hoc analysis of the BP TARGET trial assessed the effect of BPV on functional outcomes after successful EVT and showed BPV was not associated with functional outcome, corroborating our findings.

Prior studies measured BPV over a longer period of time, often up to 72 hours from stroke onset (ref), our study focused on BPV in the hyper acute phase up to 24 hours from start of embolectomy. Post hoc analysis of the CHASE (Controlling Hypertension After Severe Cerebrovascular Event) Trial found that the BPV in the acute phase was not associated with the poor outcome^12^; however, the BPV in the subacute phase of severe stroke was strongly correlated with poor outcome at 90 days (ref). Additionally, the Fukuoka Stroke Registry supported these findings by showing that in patients with AIS, the BPV in the subacute phase was independently associated with three-month poor outcomes, while no association was found in the acute phase^13^.

Additionally, while our study did not show a positive correlation between BPV and outcome, our BPV was lower than prior populations likely due to more accurate BP monitoring and control. This may also demonstrate that blood pressure in the hyper acute 24 hour phase is less variable compared with up to 72 hours. This finding may also be related to the methods used to monitor BP at academic CSCs with dedicated neurologic monitoring and post stroke care.

Given the previously demonstrated association between BPV and stroke outcomes, monitoring and controlling BPV should be considered an integral part of post-stroke care. Close surveillance of BPV can help identify patients at higher risk of poor outcomes, allowing for early intervention and tailored treatment approaches. Targeted interventions to stabilize blood pressure fluctuations and maintain optimal cerebral perfusion may have the potential to improve patient outcomes and functional recovery, as demonstrated in our patient population.

This study found no significant differences in outcome in male and female patients with good revascularization after EVT. The systems of care provided by an academic, CSC ensure that all patients’ care is expedited and provides the best opportunity for a good outcome in both sexes.

Studies of sex differences in EVT treatment and outcome in multiple stroke care settings are vital to reduce disparities in care.

## Limitations

Our study does have limitations requiring further investigation. First, there are inherent limitations in the retrospective nature of the study. Prospective assessment of BPV could provide more accurate data, but could also be at risk for for the statistical Hawthorne effect. This study was limited to two, related Comprehensive Stroke Centers. Studies in a variety of stroke center settings would provide more heterogenous data that reflects real-world clinical practice. Blood pressure was measured at three time points that were felt to be most critical in the EVT process. By limiting the data collection points, we did not capture all variability across the initial 24 hour period. Finally, only successful EVTs were examined. BPV may be even more critical in non-successful EVTs and should be examined.

## Future Directions

This study provides a foundation for the impact of intensive monitoring and expert care on reduction of BPV in a Comprehensive Stroke Center. Prospective assessment of BPV in the hyperacute phase of stroke care in this setting is vital to establishing the low BPV in this study as a class effect of CSCs or as a unique characteristic of this population or center. The comparison of BPV and outcome in PSCs, CSCs, and other stroke center settings would provide guidance on development of guidelines and best practices to reduce BPV in the overall acute stroke population. Assessment of more blood pressure datapoints throughout the hyperacute phase of AIS would also allow us to analyze the hyperacute period in which BPV drives outcome the most.

## Clinical Implications

High Blood Pressure Variability has been shown to be a factor in poor outcome in vascular disease. Reduction of blood pressure variability has the potential to preserve steady and appropriate cerebral perfusion during acute stroke. Reducing BPV may reduce the risk of sICH and improve overall outcome after stroke. Blood pressure should be monitored closely and frequently in the hyperacute stroke period and managed in a way to reduce the range of peaks and valleys in blood pressure readings in order to mimic typical cerebral autoregulation. Use of antihypertensive agents that reduce variability and allow for smooth and sustained blood pressure control in hyperacute stroke should be initiated early and titrated to achieve the goal blood pressure as quickly as possible to provide evidence based benefit.

## Data Availability

De-identied data is available upon request

## Nonstandard Abbreviations and Acronyms

BPV: Blood Pressure Variability
ARU: Acute Rehabilitation Unit
SNF: Skilled Nursing Facility
EVT: Endovascular Therapy
AIS: Acute Ischemic Stroke
mRS: modified Rankin Scale

---

**Table 1.**

| Characteristic |  |
| --- | --- |
| Mean age (years) | 70±14 |
| Sex (female) | 51.4% |
| Race |  |
| White | 49% |
| Asian | 6.3% |
| Black | 5.5% |
| Native Hawaiian Pac. Isl. | 1.6% |
| Unspecified | 37.5% |
| Hispanic ethnicity | 30.4% |
| Median NIHSS | 17±8 |
| Mean BMI | 27±8 |
| Etiology (cardioembolic) | 57.2% |
| History of HTN | 57.1% |
| History of antihypertensives | 30.5% |

